## Supplemental Appendix for "Researching COVID to enhance recovery (RECOVER) adult study protocol: Rationale, objectives, and design"

### Supporting information

#### S1 Figure: Protocol Development Timeline

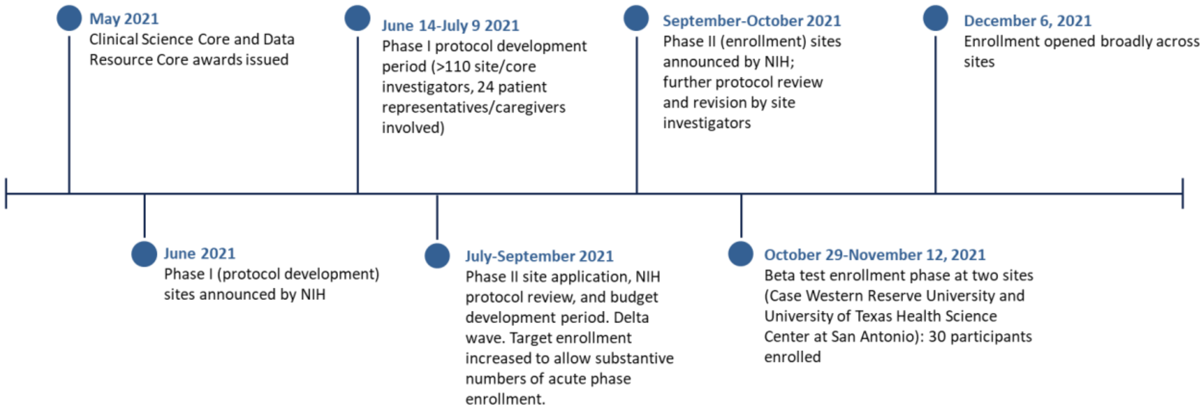

**S2 Figure: RECOVER Consortium Oversight Structure**

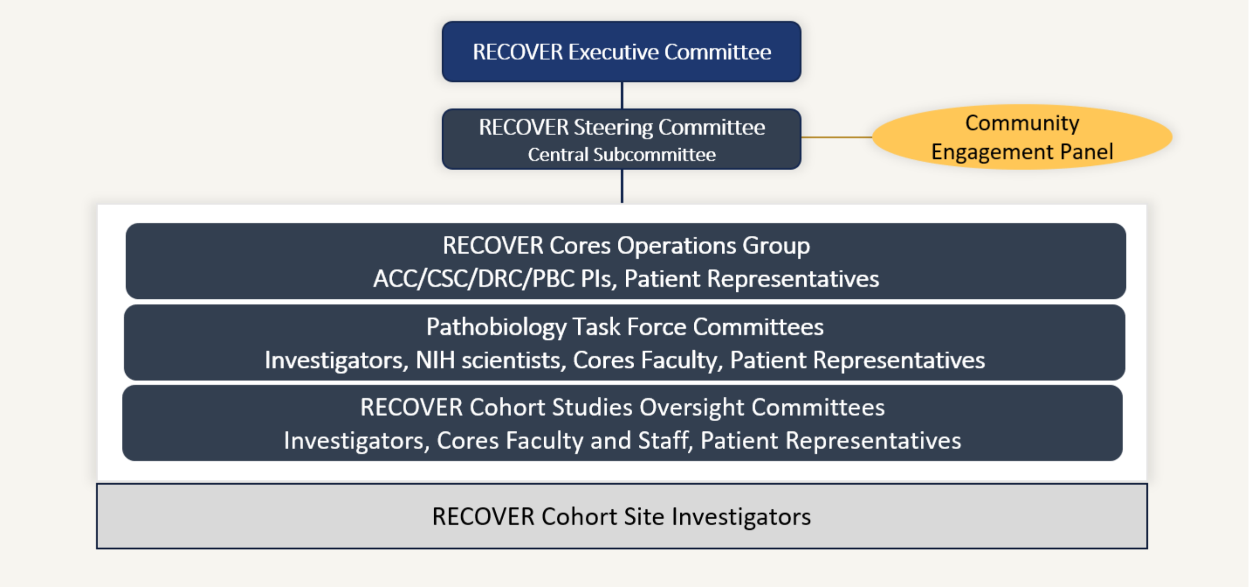


ACC: Administrative Coordinating Center; CSC: Clinical Science Core; DRC: Data Resource Core; PBC: PASC Biorepository Core; PI: Principal Investigator; NIH: National Institutes of Health

#### S1 Table: Hubs and Enrolling Sites

| Hub/consortium name | Enrolling site | Location |
|---|---|---|
| Atlanta RECOVER Clinical Research Site (CRS) | Atlanta VA Health Care System | Georgia |
| Atlanta RECOVER Clinical Research Site (CRS) | Emory Healthcare, Hope Clinic | Georgia |
| Atlanta RECOVER Clinical Research Site (CRS) | Grady Health System | Georgia |
| Atlanta RECOVER Clinical Research Site (CRS) | Kaiser Permanente of Georgia | Georgia |
| Atlanta RECOVER Clinical Research Site (CRS) | Morehouse School of Medicine | Georgia |
| Boston COVID-19 Recovery Cohort | Beth Israel Lahey Health | Massachusetts |
| Boston COVID-19 Recovery Cohort | Boston University, Boston Medical Center | Massachusetts |
| Boston COVID-19 Recovery Cohort | Brigham and Women's Hospital | Massachusetts |
| Boston COVID-19 Recovery Cohort | Cambridge Health Alliance | Massachusetts |
| Boston COVID-19 Recovery Cohort | Massachusetts General Hospital | Massachusetts |
| Boston COVID-19 Recovery Cohort | South Shore Hospital | Massachusetts |
| Boston COVID-19 Recovery Cohort | Tufts Medical Center | Massachusetts |
| Deep South SARS-CoV-2 Recovery Cohort | University Medical Center New Orleans | Louisiana |
| Deep South SARS-CoV-2 Recovery Cohort | University of Alabama at Birmingham | Alabama |
| Deep South SARS-CoV-2 Recovery Cohort | University of South Alabama | Alabama |
| Howard University | Howard University | Washington DC |
| Howard University | Mercy Medical Center, University of Maryland | Maryland |
| IDeA States Consortium for Clinical Research (ISCORE) | Hispanic Alliance for Clinical and Translational Research, University of Puerto Rico Medical Science Campus | Puerto Rico |

| Hub/consortium name | Enrolling site | Location |
|---|---|---|
| IDeA States Consortium for Clinical Research (ISCORE) | Louisiana State University Health Sciences Center New Orleans | Louisiana |
| IDeA States Consortium for Clinical Research (ISCORE) | Pennington Biomedical Research Center | Louisiana |
| IDeA States Consortium for Clinical Research (ISCORE) | MaineHealth Northern New England IDeA CTR | Maine |
| IDeA States Consortium for Clinical Research (ISCORE) | Sanford Health | South Dakota |
| IDeA States Consortium for Clinical Research (ISCORE) | Tulane School of Medicine | Louisiana |
| IDeA States Consortium for Clinical Research (ISCORE) | University of Hawaii Mountain West CTR | Hawaii |
| IDeA States Consortium for Clinical Research (ISCORE) | University of Kansas Medical Center | Kansas |
| IDeA States Consortium for Clinical Research (ISCORE) | University of Kentucky | Kentucky |
| IDeA States Consortium for Clinical Research (ISCORE) | University of Mississippi Medical Center | Mississippi |
| IDeA States Consortium for Clinical Research (ISCORE) | University of Nebraska Medical Center | Nebraska |
| IDeA States Consortium for Clinical Research (ISCORE) | University of Oklahoma Health Sciences Center | Oklahoma |
| IDeA States Consortium for Clinical Research (ISCORE) | West Virginia University | West Virginia |
| ILLInet | BrightStar Community Outreach | Illinois |
| ILLInet | Illinois Unidos | Illinois |
| ILLInet | Mile Square Health Centers | Illinois |
| ILLInet | Peoria City, County Health Department | Illinois |
| ILLInet | UI Hospital & Clinics | Illinois |
| Mount Sinai PASC Coalition (SinaiPACT) | Icahn School of Medicine at Moloraunt Sinai | New York |
| Mountain States PASC Collaborative | Bateman Horne Center | Utah |

| Hub/consortium name | Enrolling site | Location |
|---|---|---|
| Mountain States PASC Collaborative | Denver Health and Hospital Authority | Colorado |
| Mountain States PASC Collaborative | Intermountain Healthcare | Utah |
| Mountain States PASC Collaborative | University of Colorado Anschutz Medical Campus | Colorado |
| Mountain States PASC Collaborative | University of New Mexico Health Sciences Center | New Mexico |
| Mountain States PASC Collaborative | University of Utah | Utah |
| NorthEast Ohio Covid United for REcovery (NEO-CURE) | Case Western Reserve University, University Hospitals Health System | Ohio |
| NorthEast Ohio Covid United for REcovery (NEO-CURE) | Case Western Reserve University, The MetroHealth System | Ohio |
| The Pacific Northwest Consortium for Post-Acute Sequelae of SARS-CoV-2 Infection | Institute for Systems Biology | Washington |
| The Pacific Northwest Consortium for Post-Acute Sequelae of SARS-CoV-2 Infection | Providence Swedish Medical Center | Washington |
| The Pacific Northwest Consortium for Post-Acute Sequelae of SARS-CoV-2 Infection | University of Washington | Washington |
| The Pacific Northwest Consortium for Post-Acute Sequelae of SARS-CoV-2 Infection | Providence Sacred Heart Medical Center | Washington |
| The Pacific Northwest Consortium for Post-Acute Sequelae of SARS-CoV-2 Infection | Providence Regional Medical Center Everett | Washington |
| The Pacific Northwest Consortium for Post-Acute Sequelae of SARS-CoV-2 Infection | Cedars Sinai LA | California |
| PREVAIL South Texas | University of Texas Health Science Center at San Antonio | Texas |
| PREVAIL South Texas | University of Texas Education and Research Center at Laredo | Texas |
| Stanford Post-Acute Recovery Cohort (SPARC) | Stanford University | California |

| **Hub/consortium name** | **Enrolling site** | **Location** |
|---|---|---|
| Stanford Post-Acute Recovery Cohort (SPARC) | Stanford Health Care Tri-Valley | California |
| United Against COVID - AZPC3 Consortium | Banner University Medical Center Tucson | Arizona |
| United Against COVID - AZPC3 Consortium | Banner University Medical Center, Phoenix | Arizona |
| United Against COVID - AZPC3 Consortium | University of Arizona | Arizona |
| University of California San Francisco | Chan Zuckerberg San Francisco General Hospital | California |
| University of California San Francisco | San Mateo County Health Department | California |
| University of California San Francisco | UCSF Parnassus Medical Center | California |
| PRIORITY: Post-Acute Sequelae of SARS-CoV-2 in Pregnant Women and their Children | University of California San Francisco, and nationally through home visits | California |
| MFMU PASC-PREG | University of Utah (Pregnancy) | Utah |
| MFMU PASC-PREG | University of Alabama at Birmingham (pregnancy) | Alabama |
| MFMU PASC-PREG | University of Texas Health Science Center at Houston | Texas |
| MFMU PASC-PREG | University of Texas Medical Branch at Galveston | Texas |
| MFMU PASC-PREG | UH MacDonald's Women's Hospital | Ohio |
| MFMU PASC-PREG | Yale University | Connecticut |
| MFMU PASC-PREG | University of Pittsburgh, Magee | Pennsylvania |
| MFMU PASC-PREG | University of Pennsylvania | Pennsylvania |
| MFMU PASC-PREG | University of North Carolina Chapel Hill | North Carolina |
| MFMU PASC-PREG | Saint Peter's University Hospital | New Jersey |
| MFMU PASC-PREG | Ohio State University | Ohio |
| MFMU PASC-PREG | Northwestern University | Illinois |

| Hub/consortium name | Enrolling site | Location |
|---|---|---|
| MFMU PASC-PREG | NorthShore University HealthSystem | Illinois |
| MFMU PASC-PREG | New York-Presbyterian, Queens | New York |
| MFMU PASC-PREG | Miami Valley Hospital | Ohio |
| MFMU PASC-PREG | Case Western, MetroHealth Medical | Ohio |
| MFMU PASC-PREG | Medical College of Wisconsin | Wisconsin |
| MFMU PASC-PREG | Good Samaritan | Ohio |
| MFMU PASC-PREG | Duke University Medical Center | North Carolina |
| MFMU PASC-PREG | ChristianaCare | Delaware |
| MFMU PASC-PREG | Memorial City Medical Center | Texas |
| MFMU PASC-PREG | Brown University, Women & Infants Hospital of Rhode Island | Rhode Island |
| MFMU PASC-PREG | LBJ Hospital | Texas |
| MFMU PASC-PREG | Columbia University | New York |
| MFMU PASC-PREG | University of Colorado | Colorado |

**S2 Table: Survey Topics as of Protocol Version 7.0***

| Survey instrument | Topic | Asked at follow up | Source of question, if not RECOVER |
|---|---|---|---|
| Demographics | Name and contact information (retained locally) | ✓ | |
| Demographics | Alternate contacts (retained locally) | ✓ | |
| Demographics | Date of birth | | |
| Demographics | Race and ethnicity | | All of Us |
| Demographics | Biological sex | | RADx Global Codebook |
| Demographics | Gender identity | | All of Us |
| Demographics | Sexual orientation | | All of Us |
| Social determinants | Education | | NHANES |
| Social determinants | Number of people in household | | American Community Survey |
| Social determinants | Homelessness | ✓ | |
| Social determinants | Description of living place | | American Community Survey |
| Demographics | Marital status | ✓ | BRFSS |
| Social determinants | Employment | ✓ | RADx Global Codebook |
| Social determinants | Health insurance | ✓ | American Community Survey |
| Social determinants | Birthplace | | American Community Survey |
| Social determinants | Primary language | | California Health Interview Survey |
| Social determinants | Fluency in English | | California Health Interview Survey |

| Survey instrument | Topic | Asked at follow up | Source of question, if not RECOVER |
|---|---|---|---|
| Social determinants | Income in 2019 | | National Health Interview Survey |
| Social determinants | Financial insecurity | ✓ | RAND American Life Panel Impacts of COVID-19 Survey |
| Social determinants | Food insecurity | | Hunger Vital Sign |
| Social determinants | Access to health care | | National Health Interview Survey |
| Social determinants | Social support | | Medical Outcomes Study (MOS) Social Support Survey |
| ~~Social determinants~~ | ~~Loss of insurance because of COVID pandemic~~ | | |
| Social determinants | Community cohesion | | Project on Human Development in Chicago Neighborhoods |
| Social determinants | Discrimination | | Everyday Discrimination Scale |
| Social determinants | Alcohol and substance use | ✓ | TAPS Part 1 |
| Baseline disability | Baseline disability | | CDC Disability |
| Acute COVID | Diagnosis method | | |
| Acute COVID | Site and level of care for initial infection | | WHO PASC CRF |
| Acute COVID | Treatments received for initial infection | | Modified WHO PASC CRF |
| Pregnancy | Pregnancy status | ✓ | RADx-UP |
| Pregnancy | Pregnancy outcomes | ✓ | Modified WHO PASC CRF |
| Vaccination | Vaccination status and vaccine details | ✓ | WHO PASC CRF |

| Survey instrument | Topic | Asked at follow up | Source of question, if not RECOVER |
|---|---|---|---|
| Comorbidity | Immunocompromised condition and specific types | ✓ | |
| Comorbidity | Rheumatologic, autoimmune or connective tissue disease and specific types | ✓ | |
| Comorbidity | Diabetes and specific type | ✓ | |
| Comorbidity | Kidney disease and specific type | ✓ | |
| Comorbidity | Active cancer and specific type | ✓ | |
| Comorbidity | Dementia or cognitive impairment and specific type | ✓ | NIH/NINDS NeuroCOVID Databank |
| Comorbidity | Central nervous system infection, inflammatory disease or demyelinating disease and specific type | ✓ | NIH/NINDS NeuroCOVID Databank |
| Comorbidity | Seizure disorder | ✓ | |
| Comorbidity | Neuromuscular disease and specific type | ✓ | NIH/NINDS NeuroCOVID Databank |
| Comorbidity | Movement disorder and specific type | ✓ | NIH/NINDS NeuroCOVID Databank |
| Comorbidity | Cardiovascular disease and specific type | ✓ | |
| Comorbidity | Stroke or bleed and specific type | ✓ | NIH/NINDS NeuroCOVID Databank |
| Comorbidity | Asthma | ✓ | |
| Comorbidity | Chronic obstructive pulmonary disease | ✓ | |
| Comorbidity | Other chronic lung disease | ✓ | |
| Comorbidity | Use of oxygen at home | ✓ | |

| Survey instrument | Topic | Asked at follow up | Source of question, if not RECOVER |
|---|---|---|---|
| Comorbidity | Anxiety, depression or PTSD | ✓ | |
| Comorbidity | Schizophrenia or bipolar disorder | ✓ | |
| Comorbidity | Other mental health disorder | ✓ | |
| Comorbidity | Chronic liver disease | ✓ | |
| Comorbidity | Sickle cell anemia | ✓ | |
| Comorbidity | Chronic pain syndrome or fibromyalgia | ✓ | |
| Comorbidity | Myalgic encephalomyelitis/chronic fatigue syndrome | ✓ | |
| Comorbidity | POTS or other form of dysautonomia or autonomic dysfunction and specific type | ✓ | |
| Comorbidity | Obesity | ✓ | |
| Comorbidity | Polycystic ovarian syndrome | ✓ | |
| Medications | Complete medication list | ✓ | |
| Symptoms | Global health | ✓ | PROMIS-10 v1.2 |
| Symptoms | Quality of life | ✓ | PROMIS-10 v1.2 |
| Symptoms | Physical health and function, and details | ✓ | PROMIS10 v1.2, PROMIS physical function SF 4a |
| Symptoms | Mental health and thinking | ✓ | PROMIS-10 v1.2 |
| Symptoms | Social activities satisfaction and ability | ✓ | PROMIS-10 v1.2 |
| Symptoms | Anxiety, depression, irritability | ✓ | PROMIS10 v1.2 |
| Symptoms | Fatigue | ✓ | PROMIS 10 v1.2 |

| Survey instrument | Topic | Asked at follow up | Source of question, if not RECOVER |
|---|---|---|---|
| Symptoms | Pain | ✓ | PROMIS 10 v1.2 |
| Symptoms | Post-exertional malaise | ✓ | |
| Symptoms | Weakness in limbs | ✓ | WHO PASC CRF |
| Symptoms | Fever, chills, sweats or flushing | ✓ | |
| Symptoms | Loss of or change in smell or taste | ✓ | |
| Symptoms | Pain and details | ✓ | |
| Symptoms | Headache and details | ✓ | Headache Inventory Test-6 |
| Symptoms | Chest pain and details | ✓ | Seattle Angina Questionnaire |
| Symptoms | Shortness of breath or trouble breathing and details | ✓ | modified Medical Research Council scale |
| Symptoms | Cough | ✓ | |
| Symptoms | Palpitations, racing heart, arrhythmia, skipped beats | ✓ | |
| Symptoms | Swelling of lower legs and details | ✓ | |
| Symptoms | Gastrointestinal symptoms and details | ✓ | COMPASS-31 |
| Symptoms | Bladder problems and details | ✓ | COMPASS-31 |
| Symptoms | Nerve problems and details | ✓ | PROMIS physical function 4a; Neuro-QoL SF Upper extremity function v1.0; Michigan neuropathy screener |
| Symptoms | Problems thinking or concentrating and details | ✓ | Neuro-QoL Cognitive Function SF v2.0 |
| Symptoms | Problems with sleep and details | ✓ | PROMIS Sleep |

| Survey instrument | Topic | Asked at follow up | Source of question, if not RECOVER |
|---|---|---|---|
| | | | Disturbance 8a |
| Symptoms | Orthostatic symptoms and details | ✓ | COMPASS-31 |
| Symptoms | Skin color changes and details | ✓ | COMPASS-31 |
| Symptoms | Skin rash | ✓ | |
| Symptoms | Changes in sweating | ✓ | COMPASS-31 |
| Symptoms | Excessively dry eyes | ✓ | COMPASS-31 |
| Symptoms | Excessively dry mouth | ✓ | COMPASS-31 |
| Symptoms | Excessive thirst | ✓ | |
| Symptoms | Vision problems (blurry, light sensitivity, difficulty reading or focusing, floaters, flashing lights, "snow") and details | ✓ | COMPASS-31, NEI Visual Functioning Questionnaire 25 |
| Symptoms | Problems with hearing (hearing loss, ringing in ears) and details | ✓ | |
| Symptoms | Hair loss | ✓ | |
| Symptoms | Problems with teeth or gums | ✓ | |
| Symptoms | Change in menstruation or menopause and details | ✓ | |
| Symptoms | Changes in desire for, comfort with or capacity for sex | ✓ | SHOW-Q (females), UCLA Prostate Cancer Index sexual function (males) |
| Symptoms | Depression screen and assessment | ✓ | PHQ-8 |
| Symptoms | Suicidality screen and assessment | ✓ | ~~PHQ-9,~~ CSSRS suicidality screener |
| Symptoms | Anxiety screen and assessment | ✓ | Generalized Anxiety Disorder-7 |

| **Survey instrument** | **Topic** | **Asked at follow up** | **Source of question, if not RECOVER** |
|---|---|---|---|
| Symptoms | Prolonged grief screen and assessment | ✓ | Prolonged Grief-13r |
| Symptoms | Stress | ✓ | Perceived Stress Scale-4 |
| Post-COVID utilization | Hospitalization since COVID or last assessment | ✓ | |
| Post-COVID utilization | Emergency department visit since COVID or last assessment | ✓ | |

*Strikethrough text: Measure in earlier version of the protocol

**S3 Table: Tier 1 Assessments**

| Category | Assessment | In original protocol Tier 1 | In current protocol Tier 1 (v7.0) |
|---|---|---|---|
| Clinical assessment | Height, weight, body mass index | ✓ | ✓ |
| Clinical assessment | Waist circumference | ✓ | ✓ |
| Clinical assessment | Seated vital signs (blood pressure, heart rate, respiratory rate, oxygen saturation) | ✓ | ✓ |
| Clinical assessment | 30 second sit to stand | ✓ | ✓ |
| Clinical assessment | Active standing test | ✓ | ✓ |
| Clinical assessment | Wearable with continuous remote monitoring for electrocardiogram, respiratory rate, oxygenation, sleep fragmentation, actigraphy | ✓ | ✓ |
| Laboratory study | Comprehensive metabolic panel with cystatin-C | ✓ | ✓ |
| Laboratory study | Complete blood count with differential | ✓ | ✓ |
| Laboratory study | Lipid panel | ✓ | ✓ |
| Laboratory study | Hemoglobin A1c | ✓ | ✓ |
| Laboratory study | Prothrombin time, international normalized ratio, partial thromboplastin time | | ✓ |
| Laboratory study | D-dimer | | ✓ |
| Laboratory study | Troponin | | ✓ |
| Laboratory study | N-terminal pro-brain natriuretic peptide | | ✓ |
| Laboratory study | Thyroid stimulating hormone, free T4 | ✓ | ✓ |
| Laboratory study | Anti nuclear antibody | ✓ | |
| Laboratory study | Rheumatoid factor | ✓ | |

| Category | Assessment | In original protocol Tier 1 | In current protocol Tier 1 (v7.0) |
|---|---|---|---|
| Laboratory study | Anti-cyclic citrullinated peptide antibodies | ✓ | |
| Laboratory study | EBV anti early antigen IgG, viral capsid IgM, viral capsid IgG, nuclear antigen IgG | ✓ | |
| Laboratory study | 25-hydroxy vitamin D | ✓ | ✓ |
| Laboratory study | Urinalysis | ✓ | ✓ |
| Laboratory study | Urine microalbumin and creatinine | ✓ | ✓ |
| Laboratory study | hsCRP | ✓ | ✓ |
| Laboratory study | SARS-CoV-2 spike and/or nucleocapsid antibody | ✓ | ✓ |
| Laboratory study | SARS-CoV-2 NAAT | ✓ | ✓ |

**S4 Table: Tier 2 Assessments**

| Category | Assessment | In original protocol Tier 2 | In current protocol Tier 2 (v7.0) | Performed in pregnant, <3 month post-partum |
|---|---|---|---|---|
| Examination | Home sleep test | ✓ | ✓ | ✓ |
| Examination | 6 minute walk test | ✓ | ✓ | ✓ |
| Examination | Complete neurologic exam | ✓ | | ✓ |
| Examination | Neuropathy exam | | ✓ | |
| Examination | Cardiovagal innervation testing | ✓ | | ✓ |
| Examination | Rehabilitation exam | ✓ | ✓ | ✓ |
| Examination | Ears, nose, throat exam | | ✓ | ✓ |
| Examination | Mini International Neuropsychiatric Interview (MINI) | ✓ | ✓ | ✓ |
| Examination | Vision screen | ✓ | ✓ | ✓ |
| Examination | Smell Test | ✓ | ✓ | ✓ |
| Examination | NIH Toolbox oral reading recognition test age 3+ v2.0 | ✓ | ✓ | ✓ |
| Examination | NIH Toolbox picture vocabulary test age 3+ v2.0 | ✓ | ✓ | ✓ |
| Examination | NIH Toolbox auditory verbal learning test (Rey) 8+ v2.0 | ✓ | ✓ | ✓ |
| Examination | NIH Flanker inhibitory control and attention test age 12+ v2.1 | ✓ | ✓ | ✓ |
| Examination | NIH Toolbox pattern comparison processing speed test age 7+ v2.1 | ✓ | ✓ | ✓ |
| Examination | NIH Toolbox picture sequence age 7+ v2.1 | | ✓ | ✓ |

| Category | Assessment | In original protocol Tier 2 | In current protocol Tier 2 (v7.0) | Performed in pregnant, <3 month post-partum |
|---|---|---|---|---|
| Laboratory study | Procalcitonin | ✓ | | ✓ |
| Laboratory study | EBV anti early antigen IgG, viral capsid IgM, viral capsid IgG, nuclear antigen IgG | | ✓ | ✓ |
| Laboratory study | Anti-nuclear antibody | | ✓ | ✓ |
| Laboratory study | Anti-cyclic citrullinated peptide antibodies | | ✓ | ✓ |
| Laboratory study | Rheumatoid factor | | ✓ | ✓ |
| Laboratory study | Anti-dsDNA antibody | ✓ | ✓ | ✓ |
| Laboratory study | Ro antibody | ✓ | ✓ | ✓ |
| Laboratory study | La antibody | ✓ | ✓ | ✓ |
| Laboratory study | Smooth muscle antibody | ✓ | ✓ | ✓ |
| Laboratory study | RNP antibody | ✓ | ✓ | ✓ |
| Laboratory study | D-dimer | ✓ | ✓ | ✓ |
| Laboratory study | Anti-phospholipid antibody | ✓ | | |
| Laboratory study | Troponin | | ✓ | ✓ |
| Laboratory study | N-terminal pro-brain natriuretic peptide | | ✓ | ✓ |
| Laboratory study | Adrenocorticotropic hormone and morning cortisol | | ✓ | ✓ |
| Laboratory study | Hepatitis B and C testing | | ✓ | ✓ |
| Laboratory study | Parathyroid hormone | ✓ | | ✓ |
| Laboratory study | Gamma-glutamyl transferase | ✓ | | ✓ |
| Laboratory study | Cytokine panel (IL2 receptor; | ✓ | ✓ | ✓ |

| Category | Assessment | In original protocol Tier 2 | In current protocol Tier 2 (v7.0) | Performed in pregnant, <3 month post-partum |
|---|---|---|---|---|
| | IL 1beta, 2, 4-6, 8, 10, 13, 17; interferon gamma, TNF alpha) | | | |
| Laboratory study | Supine and upright catecholamine testing | ✓ | | ✓ |
| Laboratory study | ICAM-1 | ✓ | ✓ | ✓ |
| Laboratory study | Insulin | ✓ | ✓ | ✓ |
| Laboratory study | c-peptide | | | |
| Laboratory study | Oral glucose tolerance test (time points 0, 30, 60, 120 min) | ✓ | ✓ | ✓ |
| Laboratory study | Fecal WBC | ✓ | ✓ | ✓ |
| Laboratory study | Fecal SARS-CoV-2 viral load (viral RNA and/or antigen) | ✓ | | ✓ |
| Radiology | MRI brain with gadolinium | ✓ | | |
| Radiology | Volumetric non contrast chest CT (with inspiratory/expiratory scans) | ✓ | ✓ | |
| Radiology | Dual energy chest CT with contrast | | ✓ | |
| Radiology | Resting transthoracic echocardiography with strain imaging | ✓ | ✓ | ✓ |
| Radiology | Renal ultrasound | | ✓ | ✓ |
| Radiology | Fibroscan | ✓ | ✓ | ✓ |
| Radiology | Non-contrast abdominal CT | ✓ | | |
| Procedure | Electrocardiogram | ✓ | ✓ | ✓ |
| Procedure | Spirometry, resting SpO2 and | ✓ | ✓ | ✓ |

| Category | Assessment | In original protocol Tier 2 | In current protocol Tier 2 (v7.0) | Performed in pregnant, <3 month post-partum |
|---|---|---|---|---|
| | single breath diffusion capacity | | | |
| Procedure | Tilt table test | ✓ | | ✓ |

CT: computed tomography; MRI: magnetic resonance imaging

* The target window for performance of Tier 2 assessments is within 90 days of the date the study was triggered. Assessments not performed within that window are ineligible for completion; however, if the participant is still symptomatic and retriggers the assessment, or retriggers the assessment randomly, the study may then be performed within the new triggered window.

**S5 Table: Tier 3 Assessments**

| Category | Assessment | In original protocol Tier 3 | In current protocol Tier 3 (v7.0) | Performed in pregnant, <3 month post-partum |
|---|---|---|---|---|
| Examination | Michigan Neuropathy screen | ✓ | | ✓ |
| Examination | Utah Early Neuropathy scale | ✓ | | ✓ |
| Examination | Complete eye examination including optical coherence tomography | ✓ | ✓ | ✓ |
| Examination | ENT examination | ✓ | | ✓ |
| Examination | Audiometry | ✓ | ✓ | ✓ |
| Examination | Complete neurocognitive testing (to include Digit span forward/backward; Trailmaking A (3 min); Trailmaking B (3 min); Hooper Visual Organization Test, MOCA/blind MOCA, multilingual naming test, phonemic test, Benson complex figure copy, CERAD word list, Craft Story 21 Recall)+ | ✓ | ✓ | ✓ |
| Examination | Endopat testing | ✓ | ✓ | ✓ |
| Laboratory study | Serum protein immunofixation electrophoresis | ✓ | ✓ | ✓ |
| Laboratory study | Serum B12 with methylmalonic acid | ✓ | ✓ | ✓ |
| Laboratory study | CPK, aldolase, myositis | ✓ | ✓ | ✓ |

| | | | | |
|---|---|---|---|---|
| | panel | | | |
| Laboratory study | Neurofilament light chain | ✓ | ✓ | ✓ |
| Laboratory study | Fecal SARS-CoV-2 viral load (viral RNA and/or antigen) | ✓ | | ✓ |
| Laboratory study | Fecal calprotectin | ✓ | ✓ | ✓ |
| Laboratory study | Total Tau (single molecule array SIMOA) | ✓ | ✓ | ✓ |
| Laboratory study | Anti Mullerian hormone | | ✓ | ✓ |
| Radiology | MRV/MRA brain | ✓ | | ✓ |
| Radiology | MRI brain with gadolinium | | ✓ | |
| Radiology | MRI cervical spine with and without gadolinium | ✓ | | |
| Radiology | MRI thoracic spine with and without gadolinium | ✓ | | |
| Radiology | MRI lumbar spine with and without gadolinium | ✓ | | |
| Radiology | Dual-energy chest CT with contrast | ✓ | | |
| Radiology | Chest CT pulmonary angiography | ✓ | | |
| Radiology | Ventilation/perfusion scan | ✓ | | |
| Radiology | Cardiac imaging with meta-iodobenzylguanidine (mIBG)* | ✓ | ✓ | |
| Radiology | Cardiac MRI, with and without gadolinium contrast | ✓ | ✓ | |
| Radiology | Abdominal CT with contrast | ✓ | | |

| | | | | |
|---|---|---|---|---|
| Radiology | Gastric emptying study | ✓ | ✓ | |
| Procedure | Tilt table testing with supine/upright catecholamine | | ✓ | |
| Procedure | Cardiovagal innervation testing | | ✓ | ✓ |
| Procedure | Nerve conduction study | ✓ | ✓ | |
| Procedure | Electromyography | ✓ | ✓ | |
| Procedure | Skin biopsy | ✓ | ✓ | |
| Procedure | Muscle biopsy | ✓ | ✓ | |
| Procedure | Lumbar puncture | ✓ | ✓ | |
| Procedure | Facility-based sleep study | ✓ | ✓ | ✓ |
| Procedure | Full cardiopulmonary exercise testing | ✓ | ✓ | |
| Procedure | Plethysmography lung volumes | ✓ | | ✓ |
| Procedure | Bronchoscopy | ✓ | ✓ | |
| Procedure | Right heart catheterization | ✓ | ✓ | |
| Procedure | Upper endoscopy | ✓ | ✓ | |
| Procedure | Colonoscopy with or without biopsy | ✓ | ✓ | |

* The target window for performance of Tier 2 and 3 assessments is within 90 days of the date the study was triggered. Assessments not performed within that window are ineligible for completion; however, if the participant is still symptomatic and retriggers the assessment, or retriggers the assessment randomly, the study may then be performed within the new triggered window.

#### S6 Table: Writing Committee

Leora I. Horwitz
Tanayott Thaweethai
Shari B. Brosnahan
Mine S. Cicek
Megan L. Fitzgerald
Jason D. Goldman
Rachel Hess
Sally L. Hodder
Vanessa L. Jacoby
Michael R. Jordan
Jerry A. Krishnan
Adeyinka O. Laiyemo
Torri D. Metz
Lauren Nichols
Rachel E. Patzer
Anisha Sekar
Nora G. Singer
Lauren E. Stiles
Barbara S. Taylor

#### S7 Table: RECOVER-Adult Consortium Members

##### Atlanta RECOVER Clinical Research Site

**Emory University**
*Ighovwera Ofotokun, PI*
*Rachel E Patzer, PI*
Rachael Abraham
Franchesca Aguilar
Quasheba (Yasmine) Allen
Blake Anderson
Casey Beaty
Chetna Bedi
Jasmine Berry
James Douglas Bremner
Shelby Collins
Angel Craft
D'Andrea Doyle
Jess Harding
Shilpa Krishnan
Hana Lee
Jose Daniel Leon
Dong Li
Christopher Martin
Atuarra McCaslin
Miranda Montoya
Caitlin Anne Moran
Tran Nguyen
Sofia Oviedo
ReNata Shaw
Cory Sylber
Larissa J Teunis
Kehmia Titanji

**Emory Healthcare (Hope Clinic)**
*Zanthia Wiley, PI*
Arijan Ager
Mary Atha
Mary Bower
Cartia Dixon
Rebecca Fineman
Nicole Martin Franks
Natalie Gray
Ash Grimes
Evan Gutter
Lisa Harewood
Lauren Nicole Hewitt
Christopher Huerta
Brandi Johnson
Lana Khalil
Dean Kleinhenz
Athena Koumanelis
Alexandra Koumanelis
Matthew Lee
Kennedy Lewis
Matthew Litvack
Nour Makkaoui
Eileen Osinski
Bernadine Panganiban
Dilshad Rafi Ahmed
Kazi Rahman
Paulina Alejandra Rebolledo
Nadine Rouphael
Talib Sirajud-Deen
Veronica Smith
Andre Stringer
Jessica Traenkner
Kristen Unterberger
Kris Varney
Dongli Wang
Erika Wimberly
Terra Jean Winter

**Grady Health System**
*Tiffany Walker, PI*
Walter Asencios
Donchel Boone
Ke'Ara Brown-Smith
Jannah Elchommali
Jenny Elizabeth Han
Cynthia Ifejika
Vidhi Javia
Jerrod McIntyre
Christopher Toy
Alex Truong
Tamara Wesley

**Morehouse School of Medicine**
*Priscilla Pemu, PI*
Carmel Alvarez
Kelechi Carl-Igwe
Annette Dandy
Carla Holloway
Monica Juarez
Jan Morgan-Billinslea
Elizabeth Ojemakinde
Michael Prude
Ruvina Silva
Cheryl Simpson
Ashley Sylvera

**Atlanta VA Health Care System**
*Sushma Komakula Cribbs, PI*
Ghazal Ahmadi-Izad
Alicarmen Alvarez
Gustavo Capo
Erna Clyburn
Julie Costello
Xiangqin Cui
Rijalda Deovic
Jeanne Dow
Anyssa Francis
Julia Gallini
Liliana Hernandez
Ketteria Ingram
Jordi Lainez
Vincent Charles Marconi
Loice Mbogo
Abeer Moanna
Elena Morales
Yolanda Paredes-Gaitan
Chantrice Rogers
Tehquin Tanner
Kodasha M Thomas
Kartavya Vyas
Juton Winston
Cherry Wongtrakool

**Kaiser Permanente of Georgia**
*Jennifer C. Gander, PI*
Patricia Bush
Jamila Johnson
Imanii Kolailat
Monica Martinez
Roslin Nelson
Robert B Neuman
Bukkie Ojoawo
Marni Segall

##### Boston COVID-19 Recovery Cohort

-

**Brigham and Women's Hospital**
*Bruce Levy, PI*
Nya Alexander
Lindsey Baden
Shamik Bhattacharrya
Julie Buring
Li Qing Chen
Cheryl Clark
Charles Czeisler
Lauren Donahue
Elizabeth Gay
Sheila Hegde
Adetoun Okenla
Bisola Ojikutu
Daniela Lamas
Kwabena Lartey
Nomi Levy-Carrick
Maureen Macgowan
Karen Magsipoc
JoAnn Manson
Susan Redline
Elizabeth Sampson
Haley Schram
Howard Sesso
Scott Solomon
Jeffrey Sparks
Maria Sundquist
David Systrom
Vivian Thu
David Walt
George Washko
Rebecca Weiner
Maureen Whittelsey

**Massachusetts General Hospital**
*Ingrid V. Bassett, PI*
George Alba
Galit Alter
Jasneet Aneja
Francesca Caramazza
Geoffrey Chen
Lilly Fernandes
Sarah Flannery
Mabelin Garcia De La Rosa
Joseph Giacino
Leo Ginns
Marcia Goldberg
Colin Goodbred
Jennifer Haas
Stephanie Horsfall
Surabhi Iyer
Elena Jin
Michelle Jones
Boris Juelg
Diane Kanjilal
Arthur Kim
Jodi Kurtz
Gregory Lewis
Valeria Magallan
Kristin Meader
Winna Mowenn
Taing Nandi Aung
Daniel Pacella
Roy Perlis
Jonathan Rosand
Ashley Stuckwisch
Anisha Tyagi
Zachary Wallace
Dean Xerras
Danielle Zionts

**Beth Israel Deaconess Medical Center**
*Janet Mullington, PI*
Toluwanimi Ajayi
David Alsop
Esther Apraku
Michelle Beck
Diara Canton
Ai-Ris Collier
Andrea Collins
Rammy Dang
Sourbha Dani
Ramona Faris
Tamara Fong
Wilanda Gabriel
Sarju Ganatra
Monika Haack
Halle Hall
Kristine Hauser
Wendy Hori
Matcheri Keshavan
Elizabeth LaSalvia
Andrew Lewis
Chun Lim
Jason Maley
Edward Mercantonio
Murray Mittleman
Rita Monahan
Keishi Nambara
Emily Peachthong
Yuri Quintana
Uyen Rasphoumy
Marjorie Rowe
Jennifer Scott-Sutherland
Lynn Shaughnessy
Kathryn Stephenson
Jennifer Stevens
Gyongyi Szabo
Andrew Taylor
Siline Thai
Robert Thomas
Michael Vazquez
James Wareing
Huan Yang
Oscar Yang

**Boston University/Boston Medical Center**
*Jai Marathe, PI*
Tracy Battaglia
Anna Cervantes-Arslanian
Elizabeth Duffy
Naomi Hamburg
Misaki Kobayashi
George O'Connor
Fitzgerald Shepherd
Charles Williams

**Cambridge Health Alliance**
*Janice John, PI*
Pieter Cohen
Geetika Gupta
Margaret Lanca
Amberly Ticotsky

**Tufts Medical Center**
*Michael Jordan, PI*
*Honorine Ward, PI*
Rebecca Badore
Deborah Blazey-Martin
Renee Brody
Maher Ghamloush
Vidya Iyer
Laura Kogelman
Marvin Konstam

Bipin Malla
Olaniyi Ogundobede
Debra Poutsiaka
Rupali Ranade
Dulia Santos
Paul Summergrad
David Thaler
Lauren Tobias

**South Shore Hospital**
*Frank Schembri, PI*
Paola Castellotti
Carolyn McLaughlin
Justin O'Leary
Simone Wildes

#### Deep South SARS-CoV-2 Recovery Cohort

**University of Alabama at Birmingham**
*Jeanne Marrazzo, PI*
*Nathan Erdmann, mPI*
*Emily Levitan, mPI*
Donna Armstrong
Kenneth Blackwell III
Annalia Causey
Felice Cook
Julio Domingo
Conner Donahue
Maitlyn Eady
Jeffrey Edberg
Susan Ellen Binkley
Melissa Garner
Brandon Gray
Wanda Hall
Cady Hart
Bertha Hidalgo
Kaylen Holtzapfel
Alexis Jinright
Suzanne Judd
Teri Kennedy
Leigh Kirkwood
Megan Maier
Patricia McCormack
Kevin Mitchell
Aoyjai Montgomery
Juan Pablo Pilco
Leigh Powell
Rachael Shevin
Sidney Skipworth
Leah Spurgeon
Gregory Ware
Rosanne Wilson
Dana Woodruff

**University of South Alabama**
*Mark Gillespie, PI*
Noah Garcia-McClaney
Jamie Hansel
Jing Wu

**University Medical Center New Orleans**
*Jyotsna Fuloria, PI*
Paula Datri
Michael Hagensee
Cathryn Leggio
Allen Perkins
Amber Trauth
Siobhan Trotter
Alexander Van Deerlin
Sharon Weiser
Madeline Young

**Louisiana State University**
*Lucio Miele, PI*
Todd Brown
Erica Sutherland

#### Howard University

**Howard University**
*Hassan Ashktorab, PI*
*Hassan Brim, PI*
*Adeyinka Laiyemo, PI*
*Zaki Sherif, PI*
Emmanuel Baidoo
Chioniso Jakazi
Alem Mehari
Julius Ngwa
Monique Perret Gentil
Ruth Quartey
Akbar Solemani

**University of Maryland Mercy Medical Center (UMD-MC)**
*Paul Thuluvath, PI*
*Anurag Maheshwari, PI*
Mhret Alemu
Jordan Anderson
Mahak Chauhan
Sung Cho
Karli Goodman
Gandi Lanke
Ralph Lebron
Jina Ok
Polly Robarts
Somya Shesadri
Chau To
Hwan Yoo

#### IDeA States Consortium for Clinical Research (ISCORE)

**West Virginia University**
*Sally Lynn Hodder, PI*
James Bardes
Connie Cerullo
Michelle Chidester
Renee Clark
Sherri Davis
Cynthia Duda
Chad Glaze
Cami Handlan
Paige Harman
Shelly Howenstein
Karen Hunter
Shruti Jaiswal
Joy Juskowich
Wesley Kimble
Stacey Koontz
Tanya Moran
Anne Oravets
Rebecca Reece
Meghan Reeves
Zhili (Julie) Shao
Shelley Welch
Haixia Tracy Yang
Suhil Zia

**Hispanic Alliance for Clinical and Translational Research**
*Carlos A. Luciano, PI*
Aracelis Arroyo
Ileana Boneta

Daniel Casiano
Sylvia Davila
Nilda Gonzalez
Marielly Lopez
Carola Lopez-Cepero Roman
Mariela Maisonet-Alejandro
Rene Marty
Mary Mays
Yaritza Moran
Litza N Pabon Malave
Sigrid Perez Frontera
Adelma Rivera
Ana C. Sala Morales
Waleska Sanchez-Vazquez
Jorge Santana
Ruth Santos

**LSU Health Sciences Center New Orleans**
*Judd Shellito, PI*
Holli Bologna
Virginia Garrison
Erin Meyaski
Mary Meyaski-Schluter

**LSU Pennington Biomedical Research Center**
*John P. Kirwan, PI*
Kimberly Banks
Emily Rachal
Emily Bebler
Grace Bella
Alexa Bennett
Erica Bertrand
Kara Devall
Gabriela Dominguez
Amber Dragg
Angela Eldredge
Elisabeth Fontenot
Greta Fry
Bethany Gildersteeve
Sara Goff
Frank Greenway
Lauren Harrington
Erin Herrin
Lisa Jones
Erin King
Leigh Lamonica
Stephen Lee
Yejee Lee
Robert Leonhard
Jennifer Levatino
Donald Lewis
Melissa Lingle
Angrielle Lloyd
Raoul Manalac
Brian Melancon
Carla Milo
Ronald Monce
Blair Pucheu
Jennifer Rood
Stacey Roussel
Connor Sanford
Monica Santos
Crissy Sharpe
Mandy Shipp
Brooklyne Smith
Aryelle Stafford
Amy Thomassie
Celeste Waguespack
Katherine Walgamotte
Aubrey Windham

**MaineHealth/Northern New England IDeA CTR**
*Clifford J. Rosen, PI*
Abigail Arruda
Emily Berg
Anne Breggia
Kathryn Brouillette
Ivette F. Emery
Marc Flore
Lindsey Gower
Teresa Martel
Lauren Moore
Darlene Peterson
Theresa Roelke
David Seder

**Sanford Health**
*Marc D. Basson, PI*
Nicole Arbach
Lora Black
Reagan Byrum
Susan Hoover
Jessica Just
Lindsay Krause
Debra Langstraat
Allison Lutz
Sara Onnen
Vanessa Williams

**Tulane School of Medicine**
*Vivian Fonseca, PI*
Emily Callegari
Shaveeta Gupta
Elvia Haynes
Roxanne Johnson
Neda Khoshkhoo
Brian Logarbo
Michele Longo
Sofia Marquez
Roberta McDuffie
Arianna Mohiuddin
Dynte Moore
Ahona Mukherjee
Aneisha Simon
Monica Smith
Neha Upadhyay
Blaire Williams
Mei Yang

**University of Hawaii/Mountain West CTR**
*Cecilia M. Shikuma, PI*
Dominic Cheung Chow
Boonyanudh Jiyarom
Bradley Mak
Eduardo Manzano
Grace Matsuura
Cris Milne
Lorna Nagamine
Debbie Ogata-Arakaki
Rachel Ouye
Victoria Rivera
Zao Zhang

**University of Kansas Medical Center**
*Mario Castro, PI*
Charles Bengtson
Chris Bessmer
Luigi Boccardi
Jonathan Boomer
Maggie Chen
Kenton Felmlee

Theresa Howard
Jaehun Jeong
Kathy Jurius
Pamela Kemp
Christina Pantalunan
Tyler Re
Adam Ruff
Betina Senat
Leah Seymour
Leslie A. Spikes
Jennifer Troyer
Vanessa Verschelden
Vianca Williams

**University of Kentucky**
*Sidney Waldo Whiteheart, PI*
Hammodah Alfar
Suzanne Arnold
Laura Ashe
Marietta Barton-Baxter
Ryleigh Board
Suzanne Burchett
John Peyton Bush
Beth Garvy
Olivia Hage
Ellen Hartman
Darren Henderson
Melissa Hollifield
Frazier Moore
James Zachary Porterfield
Martha Sim
Ryan Weeks
Jeremy Wood

**University of Mississippi Medical Center**
*Gailen Marshall, PI*
Robert Brodell
Jamie Brown
Donielle Drakes
Vishnu Garla
Sarah Glover
Michael Hall
Thomas Hudson
Kia Jones
Christopher Moore
Rachel Morris
Utsav Nandi
Kelsey Napper
Bhagyashri D. Navaleke
Leila Seidfaraji
Shashank Shekar
John Spurzem
Amy Wigglesworth

**University of Nebraska Medical Center**
*Andrew Vasey, PI*
Amira Abdus-Salaam
Karen Blessing
Daniel Copeland
Kristi DeHaai
John D Dickinson
Irena Kovacevic
LuAnn Larson
Tracy Mathisen
Marah Miller
Kristen Nickolas
Katie Ostlund
Mary Peguero
Vaishali Phatak
Alex Reed
Maria Thurow
David E. Warren
Sara Warta
Koreen Wede
Abigail Zatkalik

**University of Oklahoma Health Sciences Center**
*Judith A. James, PI*
*Timothy VanWagoner, PI*
Cristina Gale Arriens
Amanda L. Bogie
Cathy Carmichael
Brittany Karfonta
Geneva Marshall
Tiffany Moore
Valorie Owens
James Scott
Fatima Sukhera
Timothy Walsh
Drake Williams

#### ILLInet

**University of Illinois Hospital & Clinics**
*Jerry A Krishnan, PI*
Neha Atal
Aileen Baker
Sunni Barbera
Rachel Beety
Daksh Bhargava
Andrew D. Boyd
Taylor Breiter
Elizabeth Calhoun
Ruby Camacho
Michael Carrithers
Lauren Castro
Gabrielle Cavaliere
Rashmika Chalamalla
David Chestek
Tabitha Chettupally
Judith A. Cook
Dawood Darbar
Raktima Dasgupta
Marina Del Rios
Julie DeLisa
Sai Dheeraj Illendula
Kathleen Diviak
Tara Driscoll
Mark Steven Dworkin
Angela M. Ellison
Clarie Flanigan
Meghan Fortune
Meghan Fortune Donlon
Divya Francis
Michael Freedman
Lynn Gerald
Wayne H. Giles
Bayan Hammad
Sharon Hasek
Wendy Hasse
Maryann Holtcamp
Martyna Hryniewicka
Bryan Huerta
Nina Huynh
Niha Idrees
Nahed Ismail
Akash Jain
Kyle Jennette
Shrinidhi Kadkol
Grace Kadubek
Denise Kent
Jonathan D. Klein
Lucia Large
James Lash
Michele Ledbetter
James Lee

Cinthia Leman
Jun Lu
Miriam Martinez
Cammeo Mauntel-Medici
Amy McManus
Conny Mei
Martha Menchaca
Robin J. Mermelstein
Tennessee Miller
Abeer Mohamed
David Moreno
Liam Morrissy
Naoko Muramatsu
Hugh Musick
Lourdes Norwick
Richard M. Novak
Elizabeth Ochoa-Raya
John O'Keefe
Abigail Olsen
Khushboo Patel
Nicolas Perez
Neil Pliskin
Erin Pozzolano
Bellur S. Prabhakar
Bharati Prasad
Barbara Predki
Heather M. Prendergast
John G. Quigley
Ramaswamy Ramchandran
Sarah Rappe
Ann Roach
Matthew Rowley
Gowrisree Rudraraju
Melissa Rutherfoord
Jason Scheer
Tina Schuh
Jennifer Sculley
Nancy Shapiro
Jerisha Smith-Mack
Lauren Speakman
Nancy Tartt
Angela Tobin
Ellen Uppuluri
Melissa Uribe
Terry L. Vanden Hoek
Laura Villanueva
Ceolamar Ways

**BrightStar Community Outreach**
Kathy Cullick
Pastor Chris Harris
Carl Hearne
Joani Vaughan
Erron Williams

**Chicago Urban League**
Calmetta Coleman
Lela Olds

**Envision**
Adriana Mateo
Jennifer Ramirez

**Illinois Unidos**
Lisa Aponte-Soto
Javier Arellano
Maya Diaz
Hilda Garcia
Saira Garcia
Alejandra Ibañez
Marilyn Ortiz
Cesar Rolon
Maite Zapata

**Mile Square Health Centers**
*Janet Y. Lin, PI*
Judes Fleurimont

**OSF Healthcare/St. Francis Medical Center**
*John Hafner, PI*
Isidra Baker
Jennifer Bandy
Dawn Bolliger
Praneeth Chebrolu
Jennifer Dixon
Michael Downey
Lisa Gale
Keith Hanson
Kimberly Hartwig

**Peoria City/County Health Department**
Monica Hendrickson
Seth Noland
Tracy Terlinde

**Teamwork Englewood**
Brianna Hobbs

**UI College of Medicine Peoria**
*Sarah Donohue, PI*
Sara Kelly
Jerusha Boyineni
Hannah Curry
Sherrie Edmonds
Phoebe Maholovich
Sergey Malchenko
Sarah Stewart de Ramirez
Tiffany Thompson

**UnityPoint Health**
*Samer Sader, PI*
Savannah Cranford
Terri Osmulski
Praveen Sudhindra

#### Mount Sinai PASC Coalition (SinaiPACT)

**Icahn School of Medicine at Mount Sinai**
*Alexander Charney, PI*
*Girish Nadkarni, PI*
*Juan Wisnivesky, PI*
Rawan Abdel Galil
Taj Adams
Steven Ascolillo
Esther Assenso
Emilia Bagiella
Jacqueline Becker
Kirk Campbell
Adrien Canery
Erica Cha
Esther Cheng
Dayeon Cho
Alyssa Civil
Carlos Cordon-Cardo
Chase Gornbein
Kaberi Dhar
Andrew Draheim
Jeremiah Faith
Zahi Adel Fayad
Brian Fennessy
Kaleigh Fidaleo
Sacha Gnjatic

Caroline Goldstein
Ruchir Goswami
Malika Gregory
Sabina Guliyeva
Lori Harvey-Ingram
Carol Horowitz
Rachel Jackson
Minal Kale
Matthew Hill
Jillani Kamran
Seunghee Kim-Schulze
Srisundesh Kodali
Jonathan Koganov
Amy Kontorovich
Patricia Kovatch
Jennifer Kucera
Jenny Lin
Inna Lishchenko
Martina Lopez May
Kathryn Marcon
Robert Marvin
Megan McVeety
Miriam Merad
Dara Meyer
Janice Morinigo
Khadeja Moses
Maya Nussenzweig
Cindy Osei
Tiffani Padua
Anisha Pal
Louis Pasquale
Vibhuti Patel
Farah Rahman
Michelle Ramos
Lynne Richardson
Leah Samuels
Jewelle Schofield
Joyce Serebrenik
Abdullah Serri
Nicole Simons
Sarah Tsuruo
Akosua Twumasi
Jaclyn Verity
Einab Weingarten
Alexis Whitaker
Lillian Wilkins
Michell Yee
Kimia Ziafat

**Mountain States PASC Collaborative**

**University of Utah**
*Rachel Hess, PI*
Haddy Bah
Macy Barrios
Conner Beckstead
Grayson Charlton
Dagny Donohue
Amanda Edwards
Julio Facelli
Isaac Ford
Dylan Freston
Reese Gorey
Tina Greimes
Paul Hartman
Ainsley Huffman
Alfonso Lopez
Juliemar Medina
Sophia Perez
Tristan Planelles
Jenny Powell
Annie Risenmay
Laura Scarborough
MaryBeth Scholand
Suzanne Vernon

**Denver Health and Hospital Authority**
*Edward M. Gardner, PI*
Tanner Bryan
Kaitlin Buck
Kellie Hawkins
Amy Irwin
Judy Oakes
Cole Ossian

**Intermountain Healthcare**
*Kirk Knowlton, PI*
Bailee Aguirre
Jeff Anderson
Kayelee Auer
Tami Bair
Lindsay Bosh
Lorlie Evans
Chase Garrett
Dixie Harris
Katherine Herrera
Eliotte Hirshberg
Ben Horne
James Juan
Stacey Knight
Lindsay Leither
Heather Maestas
Heidi May
Gabriel Najarian
Darija Runjaic-Ward
Scott Woller
Shyanne Zubal

**University of Colorado Anschutz Medical Campus**
*Kristine Mace Erlandson, PI*
Natasha Altman
Jill Bastman
Nick Charron
Jamie Cronin
Jesse Cwik
Elen Feuerriegel
Lani Finck
Adit Ginde
Paige Graham
Amy Harjes
David Huynh
Brandon Johnson
Sarah Elizabeth Jolley
Marjorie McIntyre
Jeffrey McKeehan
Aaron Mobley
Sarah Montoya
John Nguyen
Priscilla Pando
Kayleigh Reid
Ron J. Sokol
Shelby West

**University of New Mexico Health Sciences Center**
*Hengameh Raissy, PI*
David Archuleta
Rebecca Brito
Elyce Sheehan
Noella Garcia-Soberanez
Frederick Gentry
Eve Gronert
Michelle Sue Harkins
Debbie Lovato

Noah Martinez
Lorenzo Montoya
Alisha Parada
Davin K. Quinn
Alfredo Ramos
Irena Treacher

#### NorthEast Ohio Covid United for REcovery (NEO-CURE)

**Case Western Reserve University/University Hospital**
*Grace McComsey, PI*
Sadeer Al-Kindi
Rita Abbud
John Andrefsky
Alexis Baloi
Nicholas Boldt
Melinda Caraballo
Mary Chong
Kathryn Clark
Ann Conrad
Mary Consolo
Rebecca Curtis
Lynette Curtis
Margaret Clapp
Brian D'anza
Sarah Dawson
Kathryn DiFrancesco
Jared Durieux
Ebenezer Eteshola
Jihane Faress
Michelle Gallagher
Olivia Giddings
Paul Harris
Carla Harwell
Carla Hernandez
Shirin Iqbal
Olivia Kennedy
Danielle Labbato
Antonio Levert
Jennifer Levin
Angelica Levreault
Christian Mouchati
Caleb Mavar
Keely Newson
Princess Ogbogu
Morgan Pulling
Laura Puskas
Amanda Rivera
Michael Rodgers
Theresa Rodgers
David Rosenberg
Arnab Roy
Sarah Scott
Niyati Sheth
Beth Smith
Megan Tribout
George Yendewa
David Zhang
Sokratis Zisis

**The MetroHealth System**
*Nora Singer, PI*
Mohammed Abuzahrieh
Mirna Ayache
Emma Barnboym
Hailey Chesnick
Marissa Edminston
Larraine Gordesky
Carla Greenwood
Maricela Haghiac
Elizabeth Kaufman
Rebecca Lowenthal
Ketrin Lengu
Bridget Mackin
Shahdi Malakooti
Judy Minium
Christine Oleson
Ann Pearman
Kris Russ
Cheryl Smith
Terry Stancin
Daniel Temple
Elisheva Weinberger

#### The Pacific Northwest Consortium for Post-Acute Sequelae of SARS-CoV-2 Infection

**Institute for Systems Biology**
*James Richard Heath, PI*
Conor Brennan
Rick Edmark
Simon Evans
Vanessa Gutierrez
Jennifer Hadlock
On Ho
Kathleen Jade
Sarah Li
Andrew T. Magis
Michaela McKasson
Lee Rowen
Thea Swanson
Dan Yuan

**Cedars-Sinai Medical Center - Los Angeles, CA**
*Peter Chen, PI*
Antonina Caudill
Susan Jackman
Brittany Mattison
Sam Torbati

**Providence Regional Medical Center Everett**
*George Diaz, PI*
Angela Berrios
Jerome Differding
Vanessa Elan
Keely Heredia
Nasiha Hussain
Jo Joslin
Courtney Rinehart
Rebecca Watson

**Providence Sacred Heart Medical Center**
*Katherine Tuttle, PI*
Radica Alicic
Joni Baxter
Lisa Davis
Sarah Emerson
Claudia Flores
Susan Hood
Kelli Kuykendall
Allison Lambert

**Swedish Health Services/ISB/Swedish Medical Center**
*Jason D. Goldman, PI*
Heather Algren
Aaron Ayenew
James Del Alcazar

Allie Duven
Stephanie Johnson
John Kaneko
Christina Kim
Paula Manner
Marisa McCormack
Tija Tippett
Julie Wallick
Natalie Young

**University of Washington**
*Helen Y. Chu, PI*
Eric Chow
Nicholas Franko
Emily Guthrie
Megan Kemp
Jennifer Logue
Denise McCulloch
Dylan McDonald
Callista Nackviseth

#### PREVAIL South Texas

**University of Texas Health Science Center at San Antonio**
*Thomas F. Patterson, PI*
*Barbara S. Taylor, PI*
Reed Anderson
Azaneth Arellanes
Rose Ann Barajas
Cheryl Farner
Melinda Fischer
Mark P. Goldberg
Monica Verduzco-Gutierrez
Gabrielyd Hastings
Patricia Heard
Italia Herrera
Edgar Infante
Lisa Longoria
Hillary Johnson
Johnnie Jones
Emeka Okafor
Jan Evans Patterson
Alexis Pinones
Jennifer Potter
Brian Reeves
Irma Scholler
Sudha Seshadri
Dimpy Shah
Pankil Shah
Bridgette Soileau
Pamela Solis
Carmen Stoebner
Michael Sullivan
Robin Tragus
Joel Tsevat

**UT Education and Research Center at Laredo**
*Claudia Castillo Paredes, PI*
Stephanie Alvarado

#### Stanford Post-Acute Recovery Cohort (SPARC)

**Stanford University**
*Upinder Singh, PI*
*Paul Utz, PI*
Aubrey Adio
Neera Ahuja
Shuchi Anand
Leonard Basobas
Catherine Blish
Andra Blomkalns
Jenna Bollyky
Hector Bonilla
Athanasia Boumis
Richard Brotherton
Kimberly Clinton
Liisa Dewhurst
Vaidehi Dingankar
Julia Donahue
Jorge Doranets
Jinelle Fields
Linda Geng
Rojin Ghobadi
JaVahn Iverson
Karen Jacobson
Prasanna Jagannathan
Yasmin Jazayeri
Kathryn Jee
Jaliza Johnson
Celeste Jupiter
Ryan Kelley
Naresh Khurana
Charity Kim
Andre Kumar
Amy Kuo
Jamison Langguth
Kharma Lhamo
Martina Madrigal
Chaitasi Majmudar
Yvonne Maldonado
Lisa Maredia
Ellen O'Conor
Nicole Odenwald
Andrew T. O'Donnell
Divya Pathak
Rinoka Sato
Allyson Tayag
Crystal Ton-Nu
Mary Rithu Varkey
Anita Visweswaran
Samuel Yang

**Stanford Tri Valley**
*Minjoung Go, PI*
Christopher Jamero
Xiaolin "Kathleen" Jia
Kelly Olszewski
Orlando Quintero
Jake Scott

#### United Against COVID - AZPC3 Consortium

**University of Arizona**
*Janko Nikolich-Zugich, PI*
*Sairam Parthasarathy, PI*
Carly Deal
Rodriguez Esquival
Denise
Isabella DuPre
Kacey C. Ernst
Lunar Far
Lucia Felix
Mariana Felix
Angelica Galdamez-Avila
Amanda E Galster
Omar Gomez
Edgar Gutierrez
David T. Harris
Stefanie Harris

Adrianna Hernandez
Michael Hernandez
Maria Karnafel
Colleen Kenost
Maria Khawam
Alison Koleski
Suhr Kyle
Bonnie LaFleur
Brenda Lambert
Sicily LaRue
Ryan Lee
Kylie Lew
Karen Lutrick
Nirav Merchant
Christopher Morton
Sabrina OesterleHas
Toluwanimi Olorunnisola
Courtney L Olson
Jeanette Peralta
Iliana Perez
William (Vern) Pilling
Kristen Pogreba-Brown
Eric Reiman
Denise Rodriguez
Esquivel
Megan Rumble
T. Lee Ryan
Erick Sanchez
Maria Santa Cruz
Terry Smith
Manuel Snyder
Vignesh Subbian
Kyle Suhr
Nancy K. Sweitzer
Annie Van Den Broeke
Deanna Velarde
April Yingst

**Banner University Medical Center (BUMC) Phoenix**
*Joyce Lee-Iannotti, PI*
Lynn Autry
Sabine Borwege
Maura Carriel
Jacquelynn Copeland
Marjorie DiLise-Russo
Susan Fadden
Marilyn Glassberg
Isaias Gomez
Garrett Grischo
William Hartley
Leah Hillier
Harvey Hsu
Hira Ismail
Stephanie Iusim
Michelle James
Mrinalini Kala
Erika Kenney
Daniel Kim
Kenneth S. Knox
Melanie MacNevin
Nicole Marshell
James Michelle
Ganesh Murthy
Jami Ochoa
Fatima Palacios
Elizabeth Quijada
Elizabeth Rawnsley
Elizabeth Russo
Christina Sosa
Samuel Unzek
Sheila Vadovicky
Sharry Veres

**Banner University Medical Center (BUMC) Tucson**
*Eric M. Reiman, PI*
Maria Ambrose
Christian Bime
Joy Elizabeth Bulger
Beck
Tiffanie Cagle
Austin Derma
Margaret Drury
Damien Duran
Jose Elizondo
Rebecca Foley
Brent Gary
Claudia Gonzales
Lillian Hansen
Trina Hughes
Ayesha Javed
Elizabeth Juneman
David Lieberman
Margaret McCann
Jarrod Michael Mosier
Katie Raymer
Nikki Reed
Franz Rischard
Tanya Sandhu
Francisco Soto
Carlos Tafich-Rios
Uma Reddy
Cathleen Wilson

**Mayo Clinic Scottsdale**
Chyke Abadama
Doubeni

#### University of California San Francisco

**University of California San Francisco**
*Steven Deeks, PI*
*Dan Kelly, PI*
*Jeffrey Martin, PI*
*Michael Peluso, PI*
Khamal Anglin
Urania Argueta
Kofi Asare
Amethyst Belanger
Melissa Buitrago
Aimee Cantoran
Celina Chang Song
Alexus Clark
Nicole Del Castillo
Monika Deswal
Matthew Durstenfeld
Halle Grebe
Timothy Henrich
Rebecca Hoh
Priscilla Hsue
Beatrice Huang
Billy Huang
Rania Ibrahim
Pamela Josue
Marian Kerbleski
Raushun Kirtikar
Salman Mahboob
Sadie Munter
James Lombardo
Monica Lopez
Michael Luna
Carina Marquez
Lynn Ngo
Randy Parada
Kimberly Rhoads
Antonio Rodriguez
Alma Rodriguez Lopez
Justin Romero

Dylan Ryder
Matthew So
Viva Tai
Brandon Tran
Daisy Valdivieso
Deepshika Verma
Meghann Williams
Andhy Zamora

#### Adult Pregnancy Cohort

##### PRIORITY: Post-Acute Sequelae of SARS-CoV-2 in Pregnant Women and their Children

**University of California San Francisco**
*Vanessa Jacoby, PI*
Nyat Araya
Cinthya Arellano-Melchor
Ann Chang
Isabel De La Torre
Soujanya Gade
Estefania Guerreros
Victoria Laleau
Vanessa Monzon
Marie Salem
Maria Tolentino

##### MFMU PASC-PREG

**University of Utah**
*Torri Metz, PI*
Brynlee Buhler
Noah Carson
Jacob Draper
Kevin Duff
Marie Gibson
Denise Lamb
Amanda Nelsen
Shannon Schlater
Amber Sowles
Cassandra Vance

**Brown University (Women and Infants Hospital)**
Marshall Baez
Lisa Beati
Donna Catlow
Angelica DeMartino
Shafaq Jawed
Diana Kuhn
Haley Lefebvre
Paula Lorenzi
Jane Milano
Stephanie Nunez
Amanda O'Neill
Athena Poppas
Dwight Rouse
Janet Rousseau

**Case Western - MetroHealth Medical**
*Jennifer Bailit, PI*
*Kelly Gibson, PI*
Wendy Dalton
Brittany DeSantis
Bailey Diaz
Parmjit Gill-Jones
Melissa Kinas
Joan Lippus
Brian Mercer
Judi Minium
Abigail Pierse
LuAnn Polito
Ava Reese
Eugenia Sweet

**ChristianaCare**
*Matthew Hoffman, PI*
Caitlin Almeida
Carrie Kitto
Shannon Traczykiewicz
Ashley Vanneman

**Columbia University**
*Uma Reddy, PI*
Sabine Bousleiman
Sheica Cedano
Sara Echeverri
Megan Loffredo
Rupa Ravi
Noelia Zork

**Duke University Medical Center**
*Brenna Hughes, PI*
Nixaliz Cumba
Jennifer Ferrara
Lena Fried
Danielle Lanpher

**The George Washington University**
*Rebecca Clifton, PI*
Katia Barrett
Greg Sandoval
Steven Weiner

**Good Samaritan**
*Mounira Habli, PI*
Marta McClellan
Beth Sears

**Medical College of Wisconsin**
*Anna Palatnik, PI*
Mariana Karasti
Johanna Kessel
Christina Meyer
Eleanor Saffian

**Miami Valley Hospital**
*Samantha Wiegand, PI*
Kathleen Fennig
David McKenna
Emily Reynolds
Esther Kaye Snow
Rebecca Wirth

**New York-Presbyterian/Queens**
*Daniel Skupsk, PI*
Giorgi Kvashilava
Rosalyn Chan-Akeley
Sara Lucia Echeverri
Andrea Perez
Kelly Zhou

**NorthShore University HealthSystem**
*Beth Plunkett, PI*
Kevin Hascher
Dina Kapogiannis-Politis
Kathy Kearns
David Ouyang
Sunitha Suresh
Areebah Waseem

**Northwestern University**
*Lynn Yee, PI*
Dequana Jones
Gail Mallett
Audrey McMahon
Emily Miller
Mercedes Ramos
Trista Reynolds
Isabel Uribe

**Ohio State University**
*Maged Costantine, PI*
Anna Bartholomew
Stephanie Brindle
Barbara Cackovic
Sommer Chaney
Dawn Cline
Alyson Johnson
Baylee Klopfenstein
Huban Kutay
Devra Mast
Kayla McDaniel
Alexis Neri
Melanie Paglione
Sounali Perez
Sydney Rentsch
Jessica Russo
Taryn Summerfield
Yan Yuan
Mark Landon
Stephen Thung
Cynthia Shellhaas
Michael Cackovic
Heather Frey
Kara Rood
Patrick Schneider
Kartik Venkatesh
Courtney Abshier-Ware
Erin Cleary
Jennifer Grasch
Miranda Kiefer
Mahmoud Abdelwahab
Joe Eid
Monique McKiever
Sophia Andreatta
Christine Field
Caroline Bank
Xiao-yu Wang
Olivia Starcher

**Saint Peter's University Hospital**
*Kristy Palomares, PI*
Imene Beche
Danielle Graziano-Carrete
Clara Perez
Molly Sklios

**University of Alabama at Birmingham**
*Alan Tita, PI*
Nitin Arora
Kenneth Max Blackwell
Mariela Blair
Nicole Burrell
Lisa Dimperio
Donna Dunn
Janatha Grant
Madison Mann
Myriam Peralta
Lakia Pettibone
Jhana Plump
Jawan Struggs-Jemison

**University of Colorado**
*Camille Hoffman, PI*
Jocelyn Phipers

**University Hospitals MacDonald's Women's Hospital**
*David Hackney, PI*
Christopher Nau

**University of North Carolina - Chapel Hill**
*John Thorp, PI*
Kelly Clark
Inez Dufresne
Chelsea Grinnan
Molly Leatherland
Kathy Lloyd
Hannah Nunn
Sally Timlin

**University of Pennsylvania**
*Samuel Parry, PI*
Christina Fazio-Pizzi
Anna Filipczak
Emily Long
Meaghan McCabe
Abigail Roche
Haresh Sehdev

**University of Pittsburgh**
Jeanette Boyce
Reagan Devine
Francesca Facco
Sarah Hankle
Rachel Hines
Maura Hohn
Sharon Price
Kayli Rodgers
Marina Rushchak
Frank Sciurba
Hyagriv Simhan
Jason Styer
John Vargo
Sarah Whelan

**University of Texas HSC at Houston**
*Hector Mendez-Figueroa, PI*
Karen Castelan-Balbuena
Cynthia Edmonds
Luz Garcia
Adrienne Gross
Felecia Ortiz
Juanita Rugerio
Zina Spears
Jenifer Treadway

**University of Texas Medical Branch at Galveston**
*George Saade, PI*
Jennifer Cornwell
Luis Pacheco
Ashley Salazar
Lisa Thibodeaux
Jennifer DeVolder

**WakeMed**
*Carmen Beamon, PI*

**Yale University**
*Christian Pettker, PI*
Donna Allard
Sherrie Bitterman

Monika Lau
Jessica Leventhal
Lauren Perley
Linda Rink

**Administrative Coordinating Center at Research Triangle Institute International**

*Lisa Newman, PI*
Quinn Barnette
Patricia Ceger
Mike Enger
Katie Fain
Tonya Farris
Sean Hanlon
David Hines
Kevin Jordan
Beth Linas
Meisha Mandal
Susan Nance
Lisa Newman
Claire Quiner
Rita Sembajwe
Gwendolyn Shaw
Vanessa Thornburg
Kendall Tosco

**Clinical Science Core at NYU Langone Health**

*Rachel Gross, PI*
*Leora Horwitz, PI*
*Stuart Katz, PI*
*Andrea Troxel, PI*
Precious Akinbo
Ramona Almenana
Malate Aschalew
Lara Balick
Jasmine Briscoe
Shari Brosnahan
Alicia Chung
Stanley Cobos
Nakia Croft
Angelique Cruz Irving
Jasmin Divers
Shari Esquenazi-Karonika
Elias Febres
Catherine Freeland
Richard Gallagher
Jennifer Hossain
Neha Kansal
Tammy Kershner
Judy Kwak
Michelle F. Lamendola-Essel
Sarah Laury
Lei Lei
Janelle Linton
Max Logan
Nadia Malik
Gabrielle Maranga
Lia Mamistvalova
Maika Mitchell
Praveen C. Mudumbi
Erica Nahin
J.R. Rizzo
Johana Rosas
Chelsea Rose
Christina Saint Jean
Naomi Simon
Miranda Stinson
Mary Thomas
Lorna Thorpe
MeeLee Tom
Mmekom Udosen
Carlos Valencia
Jessica Velazquez-Perez
Crystal Vidal
Amy Willerford
Marion J. Wood
Shonna Yin
Susanna Zavlunova

**Data Repository Core at Massachusetts General Hospital**

*Andrea Foulkes, PI*
*Elizabeth Karlson, PI*
*Shawn Murphy, PI*
Shifa Ahmed
Layne Ainsworth
Marie-Abèle Bind
Caryn Boehm
Mark Bohen
Natalie Boutin
Victor Castro
James Chan
Vivian Gainer
Randy Gollub
James Kerr
Doug MacFadden
Richard Morse
Amber Nguyen
Bridget Perry
Lynn Simpson
Ravi Thadhani
Tanayott Thaweethai
Nich Wattansain
Griffin Weber

**PASC Biorepository Core at Mayo Clinic**

*Mine Cicek, PI*
Nancy Chang
Evan Ellingworth
Jordan Weyer
Jennifer Wheeler
Samantha Wirkus
Nicole Zahnle

#### S8 Table: RECOVER-Adult Committees and Task Forces

##### Executive

Hugh Auchincloss
Diana Bianchi
Joe Breen
Patti Brennan
Jeffrey Burns
Nakela Cook
Emily Cunningham
Felicia Davis Blakley
Betty Diamond
Mitchell S.V. Elkind
Tonya Farris
Lee Fleisher
Andrea Foulkes
Gary Gibbons
Laurie Gutmann
Michael Iademarco
Stuart Katz
Walter Koroshetz
Eldrin F. Lewis
Peter Marks
Hilary Marston
Mitchell Miglis
Lisa Newman
Tracy Nolen
Carlos A. Pardo-Villamizar
Amy Patterson
Sam Posner
Wendy S. Post
Serena Spudich
Clinton Wright
Heather Yates
Kanecia Zimmerman

##### Steering

Malate Aschalew
Audie Atienza
Charles Bailey
R. Graham Barr
Andra Blomkalns
Melissa Bondy
Hassan Brim
Jeffrey Burns
Alexander Charney
Benjamin Chen
Mine Cicek
John Crary
Dawood Darbar
Sean Deoni
Kathi Diviak
Ray Ebert
Jamie Elifritz
Amy Elliott
Robert L. Ferrer
John Fessel
Aloke Finn
Thomas Flotte
Andrea Foulkes
Emily Gallagher
Maria Gennaro
Rachel Gross
Melissa Haendel
James Heath
Rachel Hess
Sally Hodder
Carol Horowitz
Leora Horwitz
Vanessa Jacoby
Sarah Jolley
Suzanne Judd
Elizabeth Karlson
Stuart Katz
Rainu Kaushal
Lawrence Kleinman
Jerry Krishnan
Craig Lefebvre
Lei Lei
Emily Levitan
Bruce Levy
Daniel Liu
Jeffrey Martin
Grace McComsey
Julie McMurry
Robin J. Mermelstein
Torri Metz
Lucio Miele
Sindhu Mohandas
Janet Mullington
Shawn Murphy
Jane Newburger
Lisa Newman
Igho Ofotokun
Princess Ogbogu
Michelle Olive
Sairam Parthasarathy
Thomas Patterson
Priscilla Pemu
James (Zach) Porterfield
Antonello Punturieri
R. Ross Reichard
Jane Reusch
Kyung Rhee
Kathleen Rodgers
Juan Salazar
Lisa Schwartz-Longacre
Sudha Seshadri
Howard Sesso
Eyal Shemesh
Allan Shipp
Upinder Singh
Jessica Snowden
Serena Spudich
Cheryl Stein
Melissa Stockwell
James Stone
Jun Sun
Mehul Suthar
David Systrom
Brittany Taylor
Stephen Thibodeau
Andrea Troxel
PJ.Utz
Tiffany Walker
David Warburton
Gail Weinmann
Neely Williams
Dana Wolff-Hughes
John Wood

##### Adjudication

Khamal Anglin
Emilia Bagiella
Ryan Branski
Rodica Busui
Marissa Diggs
Vivian Gainer
Sunanda Gaur
Linda Geng
Sarah Jolley
Sarah Laury
Jai Marathe
Lisa McCorkell
Jarrod Mosier
Binita Shah
Dimpy Shah
Tiffany Walker
Peter Whitesell

**Ancillary Studies**

Hassan Ashktorab
Christine Bevc
Karyn Bishof
Yu Chen
Lori Chibnik
Dani Dumitriu
Jennifer Frontera
Paul Goepfert
Sylvie Goldman
Stephen Hewitt
Matt Huentelman
Barbara Karp
Jerry Krishnan
Marrah Lachowicz-Scroggins
Sarah Laury
Bruce Levy
Miriam Merad
Shawn Murphy
Janko Nikolich-Zugich
Laura Pace
Alice Perlowski
Brian Reeves
Juan Salazar
Sujata Thawani
Hannah Valantine
Drenna Waldrop

**Cardiopulmonary**

Natasha Altman
Soham DasGupta
Marissa Edminston
Josh Fessel
Aloke Finn
Tyler Gustafson
Francois Haddad
Jennifer Hossain
Priscilla Hsue
Pavitra Kotini-Shah
Sankaran Krishnan
Anu Lala-Trindade
Simon Lee
Alem Mehari
Patricio Millar Vernetti
Andre L Moreira
Anoop Nambiar
Robert Padera
Gail Pearson
Dhaval Raval
Franz Rischard
Erika Rosenzweig
Barbara Sampson
Frank Sciurba
Jackie Szmuszkovicz
Julie Thompson
Dongngan Truong
Viola Vaccarino
Alison Van Dyke
George Washko
John Wood

**Commonalities with Other Post-viral Syndromes**

Hector Bonilla
Christine Capone
Sekai Chideya-Chihota
Dane Cook
Walter Dehority
Monica Gutierrez
Rohan Hazra
Leonard Jason
Phillip Joseph
Dan Kelly
Joyce Lee-Ianotti
Vincent Marconi
Joshua Milner
Benjamin Natelson
Lisa O'Brien
Carlos Oliveira
James (Zach) Porterfield
Claire Quiner
Zaki Sherif
Nora Singer
Inderjit Singh
Jessica Snowden
David Systrom
C. Sabrina Tan
Emily Taylor
Vanessa Thornburg
Suzanne Vernon

**Core Operations Group**

Quinn Barnette
Frank Blancero
Mine Cicek
Lauren Decker
Jasmin Divers
Ray Ebert
Tonya Farris
Valerie Flaherman
Thomas Flotte
Andrea Foulkes
Rachel Gross
Sally Hodder
Leora Horwitz
Beth Karlson
Stuart Katz
Craig Lefebvre
Lei Lei
Shawn Murphy
Lisa Newman
Michelle Olive
Tony Punturieri
Lisa Schwartz Longacre
Upinder Singh
Stephen Thibodeau
Andrea Troxel
David Warburton
Jordan Weyer

**Health Equity/ PRO / Community Engagement**

Brett Anderson
Sujata Bardhan
Leah Castro-Baucom
Deena Chisolm
Claudia Corchado
April Joy Damian
Casey Daniel
Soham DasGupta
Walter Dehority
Candace Feldman
Josh Fessel
Lisa Goldman Rosas
Carol Horowitz
Janice John
Dhruv Khullar
Keila Lopez
Karen Lutrick
Carina Marquez
Shelly McDonald Pinkett
Larissa Myaskovsky
Lidia Regino
Kim Rhoads
Gelise St John Thomas
Sarah Stewart de Ramirez
Joel Tsevat
Carlos Valencia
Nita Vangeepuram

Anita Walden
Zanthia Wiley
Neely Williams
Shonna Yin

##### Immunology & Hematology

Hulya Bukulmez
Chris Chute
Karen Costenbader
Betty Diamond
Rao Divi
Nahed El Kassar
Nathan Erdmann
Frances Eun-Hyung Lee
Alicia Gaffney
Sacha Gnjatic
Jason Goldman
Timothy Gondre-Lewis
Jim Heath
Jennifer Hossain
Chao Jiang
Ellen Kraig
Joy Liu
Aprajita Mattoo
Joshua Milner
Sindhu Mohandas
Janko Nikolich-Zugich
Princess Ogbogu
Michael Peluso
Bellur Prabhakar
Jay Raval
Marian Sullivan
Paul Thuluvath
PJ Utz
Sidney Whiteheart

##### Integrative Physiology

Nina Caplin
Dawood Darbar
Steven Deeks
Katie Fain
Aloke Finn
Thomas Flotte
David Goldstein
Meredith Hay
Ellie Hirshberg
Charles Howell
Barbara Karp
Dean Kellogg
Rebecca Letts
Meisha Mandal
Janet Mullington
Asa Oxner
David Putrino
Jacqueline Rutter
Joel Trinity
John Wood
Roham Zamanian

##### Mechanistic Pathways

Christian Bime
Steven Bradfute
Benjamin Chen
Tom Connors
Krista Coombs
Glenn Fishman
Maria Gennaro
Timothy Henrich
Prasanna Jagganathan
Judith James
Boris Juelg
Christina Kim
Sindhu Mohandas
Michael Portman
Brian Reeves
Jalees Rehman
Ignacio Sanz

##### Metabolic Disorders

Leyna Aragon
Irina Buhimschi
Floyd (Ski)
Ralph DeFronzo
Emily Gallagher
Jennifer Hossain
Mandana Khalili
Angesom Kibreab
Tracey McLaughlin
Nandini Nair
Venkat Narayan
Elizabeth Phillips
Jane Reusch
Ivonne Schulman
Aasma Shaukat
Deborah Wexler
Jonah Zaretsky

##### Microbiology

Bill Alexander
Ami Bhatt
Hassan Brim
Shari Brosnahan
John Coffin
Adolfo Garcia-Sastre
Maria Gennaro
Joerg Graf
Timothy Henrich
Hye-Sook Him
Nahed Ismail
Jeanne Marrazzo
Rebecca McGrath
Sindhu Mohandas
Christopher Montgomery
Radu Postelincu
Ryan Ranallo
Adam Spivak
Mehul Suthar
Mary Thomas
Guangming Zhong

##### National Community Engagement Group

Teresa Akintonwa
Jasmine Briscoe
Heather Elizabeth Brown
Megan Carmilani
Marta Cerda
Debra Copeland
Felicia Davis
Roberto Garcia
Nick Guthe
Yvonka Hall
Kevin Kondo
Fadwa Lawrence
Lydia Lerma
Jacqui Lindsay
Christine Maughan
Thomas (Tony) Minor
Marjorie Roberts
Nitza Rochez
Brittany Taylor
Susanna Tranguch
Hyatt Vincent
Heather Yates

##### Neuropsychiatric

John Andrefsky
Bryan Bander
Douglas Bremner
Michael Carrithers
Melissa Cortez

Richard Gallagher
Alejandra Gonzalez
Joanna Hellmuth
Barbara Karp
Tammy Kershner
Shawn Murphy
Ganesh Murthy
Sharon H. O'Neil
Lisa Prentiss
Caitlin Rollins
Jonathan Rosand
Scott Russo
Amy Salisbury
Alan Seifert
Sudha Seshadri
Eyal Shemesh
Wendy Silver
Naomi Simon
Leanne Williams

#### Omics

Masanori Aikawa
Hassan Ashktorab
Noam Beckmann
Mike Enger
Joaquin Espinosa
Xiaowu Gai
Stephen Hewitt
Benjamin Horne
Jessica Lasky-Su
Cheryl Maier
Meisha Mandal
Lucio Miele
Emmanuel Mongodin
Lauren Nichols
Nadia Roan
Mark Russell
George Saade
Kumar Sharma
Stephanie Shiau
Jun Sun
Stephen Thibodeau
Sam Yang

#### Participant Experience

Nina Blachman
Natalie Boutin
Phoebe Burton
Marina Catallozzi
Cheryl Clark
Beth Dworetzky
Belinda Edwards
Robert L. Ferrer
Beatrice Huang
Suzanne Judd
Sarah Laury
Hugh Musick
Divya Pathak
Kristen Pogreba-Brown
Hengameh Raissy
Lynne Richardson
Russell Rothman
Laura Wagner
Ann Wallace

#### Population Science

Paul Barach
Melissa Bondy
Victor Castro
Mine Cicek
Joanne Elena
Kacey Ernst
Josh Fessel
Dan Fort
Brian Hendricks
Bertha Hidalgo
Cory Hussain
Carmen Isasi
Dan Kelly
Adeyinka Laiyemo
Margaret Lanca
Juan Lewis
Beth Linas
Heidi May
Kimberly McHugh
Naoko Muramatsu
Girish Nadkarni
Susan Nance
Kian Nguyen
Priscilla Pemu
Lisa Postow
Suchitra Rao
Dimpy Shah
Sidd Shenoy
Stephanie Wilson
Dana Wolff-Hughes

#### Presentations and Publications Oversight

Ingrid Bassett
Diana Berrent
Andra Blomkalns
Hassan Brim
Rebecca Clifton
Nathan Erdmann
Kristine Erlandson
Josh Fessel
Valerie Flaherman
Margot Gage
Mark Goldberg
Edmond Kabagembe
Tammy Kershner
Patricia Kinser
Jonathan Klein
Gregory Laynor
Grace Lee
Grace McComsey
Brian McCrindle
Julie McMurry
Girish Nadkarni
Priscilla Pemu
Dustin Rabideau
Erika Rosenzweig
David Warburton

#### QA/QC Data Integrity

Audie Atienza
Charlie Bailey
James Chan
Mine Cicek
Hannah Davis
Kathi Diviak
Ray Ebert
Dan Fort
Jennifer Gander
Janos Hajagos
Kellie Hawkins
Shahidul Islam
Dan Kelly
Tammy Kershner
Patricia Kovatch
Michelle Lamendola-Essel
Simon Li
Daniel Liu
Holden Maecker
Emily Pfaff
Anisha Sekar
Zaki Sherif
Vignesh Subbian
Anand Viswanathan
Jennifer Wheeler
Meredith Zozus

**Study Design**

Yvette Burgos
Chris Chute
Katharine Clouser
Juan Espinoza
Megan Fitzgerald
Valerie Flaherman
Elizabeth Karlson
Barbara Karp
Lawrence Kleinman
Adeyinka Laiyemo
Emily Levitan
Gabrielle Maranga
Gailen Marshall
Jeffrey Martin
Robin Mermelstein
Torri Metz
Sindu Mohandas
Jennifer Muszynski
Jane Newburger
Igho Ofotokun
Sairam Parthasarathy
Kyung Rhee
Lumy Sawaki-Adams
Mary Beth Scholand
Howard Sesso
Nora Singer
Jessica Snowden
Cheryl Stein
Lauren Stiles
Melissa Stockwell
Kelan Tantisira
Barbara Taylor
Tanayott Thaweethai
Juan Wisnivesky
